# An Exploration of the Mental Health impact among Menopausal Women: The MARIE Project Protocol (International Arm)

**DOI:** 10.1101/2023.11.26.23299012

**Authors:** Gayathri Delanerolle, Heitor Cavalini, Julie Taylor, Sharron Hinchliff, Vikram Talaulikar, Zukiswa Zingela, Teck-Hock Toh, Xiu-Sing Wong, Kim-Yen Lee, Jeffrey Soon-Yit Lee, Sanghamitra Pati, Nirmala Rathnayake, Thamudi Sundarappeuma, Tharanga Mudalige, Lanka Dasanayake, Damayanthi Dasanayake, Vindya Pathiraja, Prasanna Herath, Om Kurmi, Ashish Shetty, Muhammed Irfan, Rabia Kareem, Helen Kemp, Subrata Kumar Palo, George Uchenna Eleje, Bellington Vwalika, Wenjing Zhao, Jian Qing Shi, Peter Phiri

**Author notes:** **Corresponding author** Dr Gayathri Delanerolle, Senior Researcher, Research & Innovation Department, Southern Health NHS Foundation Trust, Clinical Trials Facility, Tom Rudd Unit Moorgreen Hospital, Botley Road, West End, Southampton SO30 3JB, UK. Shared last author (Jian Qing Shi, Peter Phiri). Shared second author (Heitor Cavalini, Julie Taylor, Teck-Hock Toh).

## Abstract

**Introduction:** Menopause is characterised by the ending of the menstrual cycle as part of a natural process. However, menopause can also be caused by other health conditions, such as premature ovarian failure or cancers that may have led to an oophorectomy or a radical hysterectomy. The physiological and psychological mechanisms linked to menopause across all age groups, races and ethnicities are not well understood. The paucity of data could reduce the advancement of optimal clinical practice, leading to reduced quality of life for women. To better explore and assess menopause, we have designed the **MenopAuse mental hEalth Rating (MARiE)** tool.

**Methods:** We will conduct a prospective mixed methods study using two workstreams of WP2a and WP2b among in women and trans-men ≥18 years old that are experiencing perimenopause, menopause or post-menopause among an array of ethnicities and races. WP2a will involve a number of validated clinical assessments of Hospital Anxiety and Depression Scale, Insomnia Severity Index Scale, Menopause Rating Scale, Greene Climacteric Scale, Health Related Quality of Life, Quebec Pain Disability Scale, and Burnout Assessment Tool will be administered digitally using the Qualtrex platform. WP2b will assess the feasibility of using a novel tool called MARiE to report face validity and efficacy.

**Ethics approval:** Research Ethics approval reference for this study in the UK is 22/EE/0159. As required, country-specific approvals have been obtained and continue to be secured.

**Dissemination:** The study findings will be made available using a peer-review publication journal, workshops and conferences.

## INTRODUCTION

Menopause is the cessation of menstruation, and the transition into this phase is characterised as the period spanning from the onset of alterations in the menstrual cycle or the emergence of vasomotor symptoms over 12 months following the last menstrual period^1^. The menopausal transition is distinguished by hormonal fluctuations and is linked to a diverse array of physiological and psychological symptoms that could lead to comorbidities such as cardiometabolic diseases or mental health conditions^1,2^. The nature and severity of these issues can significantly differ across women and trans-men, which could have a bidirectional relationship with societal factors such as cultural practices across various ethnicities, races and geographical regions^2,3^. Typically, women begin to experience these menopausal symptoms in their late 40s, and they can persist for a duration of 5 to 8 years, but in some cases, they may extend over the long term. This is referred to as natural menopause. Although, surgical or medical menopause could impact people at any age, as it is a direct result of an oophorectomy or a hysterectomy due to conditions such as cervical cancer, endometriosis or uterine fibroids^2-4^.

The Nuffield Health Survey, which encompassed 3,725 menopausal women in the UK, revealed that 47% of these women reported experiencing symptoms of depression, while one-third of them encountered feelings of anxiety^4,5^. Additionally, sleep disturbances, encompassing difficulties in falling asleep and recurrent awakenings, affect up to one-third of women going through the menopausal transition, which could further exacerbate their mental health issues. It is estimated that approximately 20% of women in this transition phase seek medical help from their primary care physicians due to symptoms of depression or anxiety, underscoring the seriousness and impact of this issue. The Study of Women’s Health Across the Nation (SWAN) has identified various contributing factors that make women more susceptible to depression during this period^5^. These factors include discomfort related to physical symptoms such as hot flashes, pre-existing mental health conditions, a lack of adequate social support, psychosocial stressors, health and lifestyle behaviours, and demographic characteristics. However, the increased risk of depression and anxiety observed in this population cannot be fully explained by these factors alone^6,7^. Evidence from studies that adjust for these variables indicates a lingering, elevated risk. It is becoming increasingly evident that the clinical manifestation of depression and anxiety in women undergoing the menopausal transition differs from that in women not experiencing these biological changes^7^. This phenomenon is associated with a wide spectrum of symptoms, including fatigue, diminished energy levels, low self-esteem, feelings of isolation, cognitive impairment, reduced libido, weight gain, muscle aches, and back pain^8^. Mood changes observed in women during the menopausal transition are predominantly characterised by irritability, paranoia, and anger, as opposed to women of reproductive age, who typically exhibit sadness and a low mood. The causation of mental health issues during the menopausal transition is multifaceted, encompassing biological, psychological, and social factors^7,8,9^. Hormonal fluctuations in the hypothalamic-pituitary-gonadal axis directly impact brain function. Gonadal hormones play a significant role in regulating brain metabolism, enhancing cerebral blood flow, reducing inflammation, and promoting neuronal regeneration and nerve growth factor^10,12^. These biological factors interact with psychosocial elements during this life stage, including perceptions of ageing, the cessation of reproductive capabilities, changing roles in the workplace and family, physical ailments, and stressful life events^11 -14^. The available evidence strongly reinforces the idea that mental health issues during the menopausal transition have a distinct origin and display specific clinical characteristics^15^. This underscores the necessity for a specialised tool designed to investigate and assess the impact of menopause on both the physical and psychological health during this phase.

## METHODS

### Aims and Objectives

This project aims to investigate the mental health impact of menopause by evaluating both the physiological and psychological aspects through several distinct workstream packages. This protocol focuses on Work Package (WP) 2, which consists of two components: work packages WP2a and WP2b. The primary objective of WP 2 is to comprehensively explore menopause across diverse populations and to further validate our novel menopause assessment tool known as the MenopAuse mental hEalth Rating (MARiE). This tool was developed by synthesising existing research data, clinical expertise, and collaborative efforts as part of the broader women’s health program, ELEMI.

### Study Design

The MARIE project uses a multifaceted approach where WP2a uses a prospective mixed-methods study design aimed at gathering additional information using existing clinically valid questions. Although these are generic and part of clinical consultations, they remain non-specific to the context of menopause. The administration of these questions will be followed by a study-specific topics guide developed to assess participant experience to further strengthen the understanding and evaluation of menopause.

### Data collection

Participants will complete the questions online along with demographic questions. The overall questionnaire includes the Hospital Anxiety and Depression Scale (HADS), Insomnia Severity Index Scale (ISIS), Menopause Rating Scale (MRS), Greene Climacteric Scale (GCS), Health-related Quality of Life (HRQoL), Quebec Pain Disability Scale (QPDS), Marital Satisfaction Scale (MSS) and the Burnout Assessment Tool (BAT-12). These questionnaires will be completed at baseline (days 1) and on day-30 following informed consent. A sub-set of participants will complete a qualitative interview on day-60 using the topics guide.

The qualitative interviews will be audio-recorded and transcribed by the local teams. Early interviews will be reviewed by the research team to assess whether any modifications to the topic guides are necessary. The anticipated interviews duration is approximately 45 minutes, although the duration may vary for each participant. The researchers will transcribe the interviews, and the transcripts will be reviewed for accuracy. Subsequently, they will be de-identified and uploaded into NVivo software version 11.2 for data coding and retrieval.

WP2b will commence as a feasibility study following the completion of WP2a. Work Package 2b is focused on validating the MARiE assessment tool in a broader participant population.

### Study population & recruitment

This study protocol will be used world-wide with a number of countries being onboarded, including but not limited to India, China, Sri Lanka, Malaysia, Zambia, South-Africa, Nigeria and Pakistan. Some aspects of the data collection will be aligned with the local requirements and cultural practices.

The eligibility criteria for WP2a and WP2b include women and trans-men aged 18 years or older and have either undergone natural or surgical menopause or they are currently undergoing the peri-menopausal or post-menopausal phase.

Participants should be willing and able to provide written consent and have access to a digital device to complete the online questionnaires and interviews. Recruitment efforts will be extended to reach potential participants through various social media platforms, including Twitter, LinkedIn, National Health Service (NHS) hospital websites, Instagram, and Facebook. Participants will also be recruited through menopausal and family physician clinics by local clinical research teams. As this study is open to rural and urban principalities, participants will receive material in their preferred local languages.

### Informed consent

Informed consent will be obtained electronically through the Qualtrics XM platform for WP2a and WP2b or manually on paper copy. Participants will receive comprehensive information about the study. Eligible individuals who provide their consent will then be officially enrolled in the study. Participants will have the option to withdraw from the study at any time before submitting their questionnaire. However, once the questionnaire is submitted, it will not be feasible to identify the participant for data withdrawal, unless they have previously provided contact details that can be used to identify and withdraw their data. In such cases, any data that has already been collected with the participant’s consent will be retained, but no additional data will be gathered, and no further research procedures will be conducted about that specific participant.

### Data analysis

Quantitative data will be collected through online questionnaires utilising the Qualtrics XM platform. Each participant will be assigned a study ID; no personally identifiable information will be recorded. Data will be extracted from the Qualtrics XM platform and imported into statistical software packages like SPSS and STATA for graphical representation and analysis. Data collection and analysis will be intertwined. If data saturation is reached, the interview focus may shift to other groups to enable more comprehensive exploration where appropriate.

WP2a: We will present descriptive statistics for continuous variables based on the data’s distribution, using means (SD) or medians (IQR) as appropriate. For categorical data, we will provide statistics in the form of frequencies and proportions. In cases where data does not follow a normal distribution, non-parametric techniques such as the Kruskal-Wallis test and Mann-Whitney U-test will be employed. Additionally, for categorical variables and to explore associations between demographic data and responses related to mental health and well-being, we will use either the Chi-square or Fisher’s exact test. Pearson’s correlation analysis will be applied to measure the strength of linear relationships between variables.

WP2b: Pearson’s correlation analysis will be used to assess the strength of linear relationships between variables. We will also evaluate the goodness-of-fit of latent class models for categorical responses through Pearson and likelihood-ratio chi-squared tests. Following this, we will employ a factor model, using confirmatory factor analysis with the Maximum Likelihood approach, to explore the covariance fit of the tested factor models. Model selection will be based on goodness-of-fit indices (GOF), considering various indices, both absolute and relative fit. All interviews will be conducted via a secure online platform, specifically a password-protected Zoom teleconference. These interviews will be audio-recorded and transcribed in their entirety. Early interviews will undergo review by the research team to determine if any adjustments to the topic guides are necessary. Data collection and analysis will be conducted in an integrated manner, employing a framework methodology that utilizes an indexing coding approach to organise the data and facilitate interpretation.

## AUTHOR’S CONTRIBUTIONS

GD conceptualised and developed the MARiE project as part of the ELEMI program. GD and JQS designed the statistical analysis plan. GD wrote the first draft of the protocol manuscript. All authors critically appraised and commented on the protocol manuscript. All authors read and approved the final manuscript.

## DATA AVAILABILITY STATEMENT

The authors will consider sharing the dataset gathered upon receipt of reasonable requests.

## List of abbreviations

MARiE: Menopause mental health Rating
GOF: Goodness of Fit

